# Initial respiratory support outcomes and associated factors among preterm neonates with respiratory distress syndrome admitted at Moi Teaching and Referral hospital Eldoret, Kenya

**DOI:** 10.1101/2024.03.17.24304436

**Authors:** Joyce Kalekye Ndeto, Winstone M Nyandiko, Audrey K Chepkemoi, Ann W Mwangi

## Abstract

**Background:** Respiratory distress syndrome (RDS) is the single most important cause of morbidity and mortality in preterm neonates. Early management of RDS is crucial in determining short- and long-term outcomes and studies have established initial respiratory support (IRS) among other factors as an important determinant. Despite preexisting guidelines and advancements in the management of RDS, IRS failure with noninvasive ventilation is common and is associated with unfavorable outcomes.

**Objective:** This study evaluated the non-invasive initial respiratory support outcomes and associated factors among preterm neonates with respiratory distress syndrome admitted in the newborn unit (NBU) at Moi Teaching and Referral Hospital Eldoret (MTRH), Kenya

**Methods:** Using a prospective observational hospital-based study, preterm neonates ≤ 35weeks admitted at the newborn unit with clinical RDS were followed up for 28 days. The primary outcome was IRS success or failure characterized by not stepping up or stepping up the respiratory support respectively within 72 hours of life and associated maternal and neonatal factors. Descriptive statistics was described using mean ± (SD) for continuous variables and frequencies and percentages for categorical variables. Simple and multinomial regression analysis was performed to evaluate relationship between different IRS methods with outcome variables and a p-value of < 0.05 was considered significant.

**Results:** We enrolled 320 neonates, 172(53.8%) were male with a mean (SD) gestation age of 30.9 (2.95) weeks. The mothers mean age was 27 years, ranging (15–43). 70(22.4%) 95%CI:17.95,27.47] had IRS failure and 243(77.6%) had IRS success. On multivariate analysis IRS success was associated with primiparity (AOR=2.81;95%CI: 1.42, 7.99), birthweight > 1300g (AOR= 5.04;95%CI 1.81, 14.6), low modified Downes score (AOR=26.395%CI 3.37, 230) and normal admission temperatures (AOR=0.32;95%CI 0.12, 0.72) (p= <0.001).

**Conclusion:** Noninvasive ventilation had a high initial respiratory support success. Primiparity, birthweight >1300g, normal admission temperatures and low Downes score were associated with IRS success.

## Introduction

Respiratory distress syndrome formerly hyaline membrane disease is a breathing disorder that occurs almost exclusively among preterm neonates. As a result of immature lungs producing inadequate surfactant levels, this leads to an increase in surface tension that will cause atelectasis. (1)

Globally, the infant mortality rate has declined significantly but neonatal mortality rates are declining at a slower pace. (2). Neonatal mortality in Kenya is at 21 deaths per 1,000 live births, with a third of those attributable to prematurity (3) RDS is the single most important cause of morbidity and mortality in preterm neonates (4) In Moi Teaching and Referral Hospital (MTRH) 37.1% and 64.6% of neonates were admitted secondary to RDS in 2006 and 2013 respectively (5)(6) and the case fatality from RDS in MTRH in 2016 was 72.3%, 61% in the first 10 days of life (7)

Management of RDS is the main determinant in both short- and long-term outcomes in neonates. Respiratory management of preterm infants with or at risk of RDS aims to maximize survival while minimizing potential adverse effects, such as bronchopulmonary dysplasia (BPD). Therefore, early identification of those premature infants who most likely fail noninvasive (NIV) can allow specific therapeutic interventions to be aimed at them.

Over time, studies on the different IRS modalities and outcomes have been carried out with the aim of establishing superiority of the different techniques. Even with these recent advances in the perinatal management of neonatal respiratory distress syndrome (RDS), the early respiratory management is still controversial with no ideal approach fully established. The rate and timing of the different modalities varies widely, apparently unrelated to severity of illness and the impact of this variability in practice is unknown.(8)

A survey done by Parat et al on respiratory management of ELBW neonates among neonatal specialist, concluded that variability in treatment strategies of RDS in ELBW is common among the specialists. (9) In lieu of this, two internationally recognized bodies, European Society for Paediatric Research and the American Academy of Pediatricians (AAP) through the European Consensus guidelines on the management of RDS 2019 update and policy statement on respiratory support in preterm infants at birth 2014 respectively, recommend use of CPAP immediately after birth with subsequent selective surfactant administration. (10)(11)

Even with these guidelines recommending CPAP as the primary mode of respiratory support, some studies have shown an increase in CPAP failure rates and poor prognostic outcomes.(12)(13) (14) (15).

Several variables have been associated with the outcomes and further understanding has led to formulation of predictive models that can guide in determining the appropriate IRS. A study by Afjeh et al showed low gestational age and birth weight and maternal disease were associated with IRS failure. (15). Another study by Kakkilaya et al, which aimed to develop a prediction model to identify infants admitted on continuous positive airway pressure (CPAP) requiring intubation within seventy-two hours of life (HOL), lower antenatal steroid exposure, birth weight, higher radiographic severe respiratory distress syndrome (RDS) and fraction of inspired oxygen (FiO2) were identified as predictors of outcome.(16).

No analogous study has been carried out since the newly established well equipped NICU at MTRH. Therefore, this study determined the current IRS outcomes and factors associated with success and failure of the different strategies. This will help guide the choice of primary respiratory support strategy and allow early identification of those preterm neonates at risk of failing NIV IRS for establishment of specific therapeutic interventions among pre-term neonates with RDS.

## Methods

This was a prospective observational hospital-based study. Preterm neonates ≤ 35 weeks gestational age were recruited within the study period and followed up to 28 days post-delivery, hospital discharge, or Death, whichever came first. The study was conducted at the Newborn Unit at the Riley Mother and Baby Hospital in Moi Teaching and Referral Hospital (MTRH) in Eldoret-Kenya. The facility is the second largest National Teaching and Referral Hospital (Level 6 Public Hospital) located in Uasin Gishu County in the North Rift region of Western Kenya. The hospital has an estimate of up to 14,000 deliveries per year. The NBU has a 60-bed capacity although on average has approximately 80-110 neonates per day. It consists of a 16-bed Neonatal Intensive Care Unit (NICU) with 16 ventilators with both invasive and non-invasive modes of ventilation. The unit also has variable flow CPAP machines with NCPAP, NIPPV, and HHHNC modes.

This study received ethical approval from the Institutional Review and Ethics Committee of Moi Teaching and Referral Hospital / Moi University School of Medicine (IREC FAN/0004043). Additionally, an administrative approval was obtained from the Chief Executive Officer (CEO) of Moi Teaching and Referral Hospital (Ref. ELD/MTRH/R&P/10/2/V.2/2010). Written informed consent was obtained from the parents of neonates (S1 File).

Fischer’s formular was used for sample size estimation, using p value of 88% based on a study by Ammari and collaborates and consecutive sampling technique was used to recruit a minimum sample size of 320. Gestational age assignment was based on 1^st^ trimester ultrasound or Ballard assessment postnatally in the absence of a first trimester ultrasound. Those above 35 weeks GA were excluded from the study due to the lower incidence of RDS among them and the higher risk of other comorbidities associated with respiratory distress. Neonates with an APGAR score of < 5 in the first 5 minutes and those with cardiopulmonary congenital anomalies were excluded because they present with symptoms of respiratory distress that overlap with those of RDS. Those with congenital disorders not compatible with life were also excluded.

Preterm neonates being admitted at the NBU with a clinical diagnosis of RDS were identified by the PI. The neonates who met inclusion criteria, the neonates’ mother or legal representative in the absence of the mother was approached for written informed consent by either the trained research assistants or the PI. After signing the informed consent, either the PI or trained research assistant began collecting the maternal and neonatal data using a semi-structured data collection form (S2 File) from the mother or legal representative and admission notes. The pediatric resident, primary pediatrician, or neonatal specialist decided on the method of respiratory support after the initial stabilization. Decisions were mainly based on the respiratory status and predesigned protocols (S3 File). We then divided the recruited preterm neonates into two groups according to the respiratory support initiated within the first 2 hours of life (17). The two classes of IRS were Group I-Noninvasive ventilation and Group II - Invasive ventilation. Group I comprised of two categories; 1- Low flow nasal canulae, High flow nasal cannula and Oxygen via a non-rebreather mask (NRM) 2- Nasal continuous positive airway pressure (NCPAP) and Nasal intermittent positive pressure ventilation (NIPPV). Those who started on delivery room CPAP were included in the study. The PI and trained research assistants followed up the neonates for 72 hours.

Within the 72-hour period, neonates in group I were placed either in the “Success: Never Upgrading” group, if there was no need for IRS improvement, or in the “Failure: Upgrading” group in case of IRS failure. Group II clinical course was followed up until extubation. The study procedure can be summarized as shown in fig 1 below.

**Figure 1.**
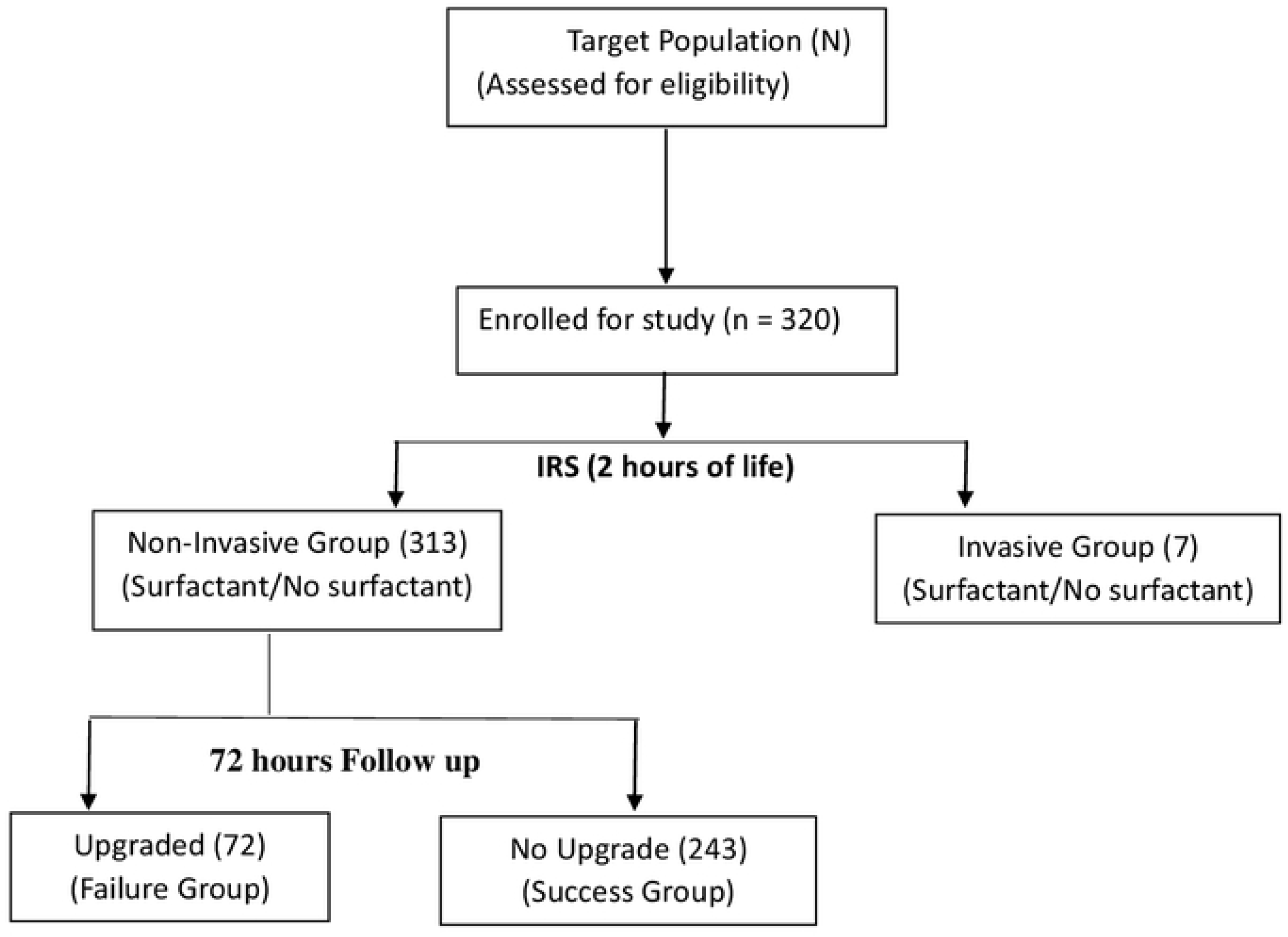

SPSS v26 was used for the statistical evaluation and Rcore2020 version 4.2.2 to run regressions. Mean and standard deviation were used to summarize continuous variables and frequencies and corresponding percentages were used to summarize categorical variables. Continuous variables were compared using independent sample t-test or Kruskal wallis test categorical variables were compared using Pearson’s Chi-Square tests or Fisher’s Exact Test (critical value ≤0.05) and odds ratios (95% confidence interval). Simple and multinomial regression analysis was used to establish relationship between different IRS methods with outcome variables. Predictor variables with *p*-value of <0.2 on bivariate analysis were included in multivariate logistics regression and a *p*-value <0.05 and calculated adjusted odds ratio (95% CI) was considered statistically significant.

## Results

This study enrolled 320 neonates, of whom 172(53.8%) were males with a mean GA (SD) of 30.6(2.95). The mean [SD] birth weight of the neonates was 1414[411] grams, ranging from 575– 2435 grams. The majority were delivered in hospital, 292(91.2%), SVD deliveries were 176(55%), followed by CS who were 125(39.1%), and the least was SBD 5.9% of the total deliveries. Singleton pregnancies were the majority at 219(68.4%) and twin pregnancies were 85(26.6%). The mean (SD) admission temperature was 35.8 (0.9) degrees Celsius, and the mean (SD) modified Downes score performed at delivery was 3.8 (1.537), ranging (1–8). As shown in Table 1 164(51.3%) had moderate respiratory distress, 147(45.9%) had mild respiratory distress and only nine had impending respiratory failure. As demonstrated in Fig 2

**Figure 2.**
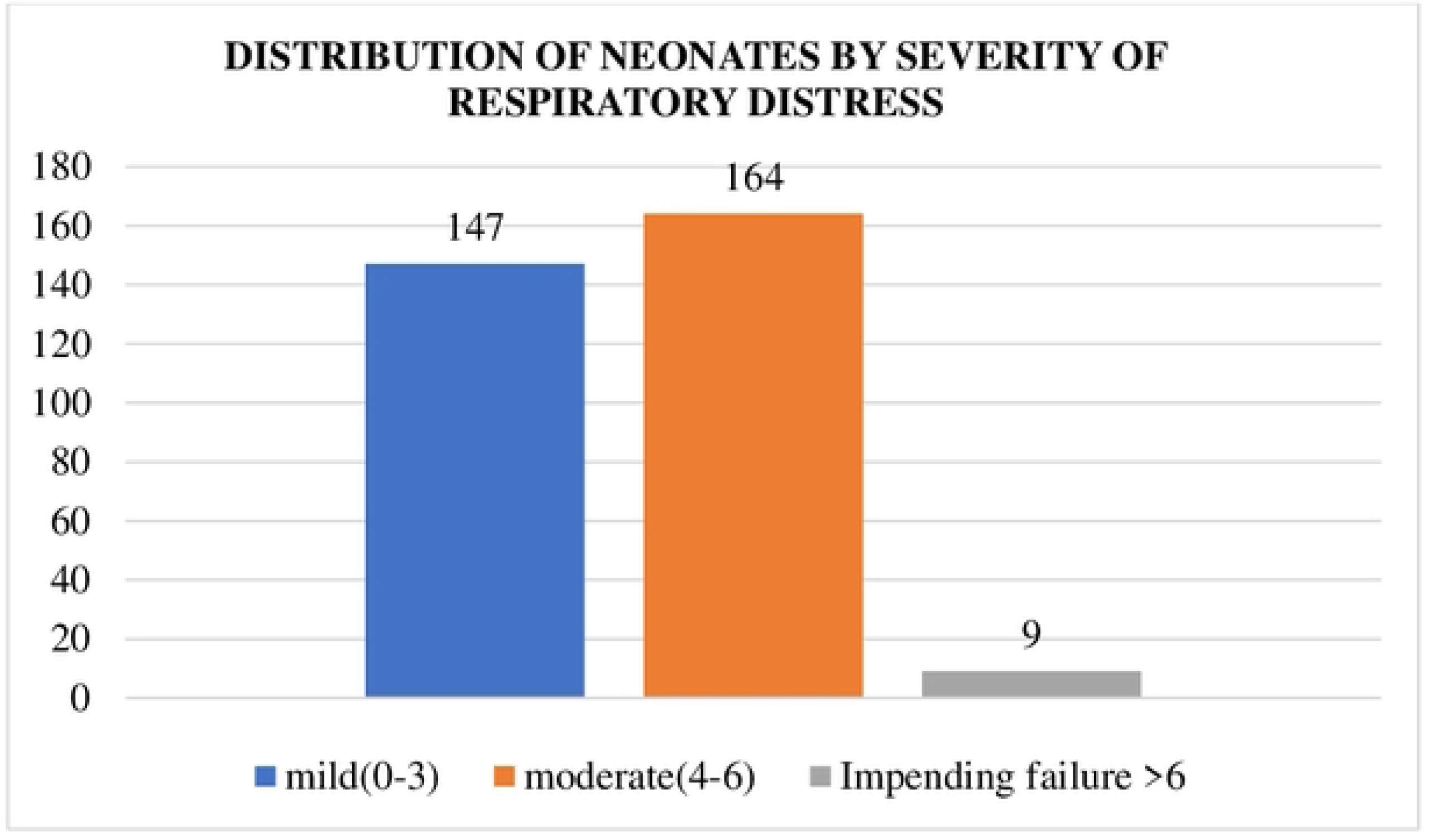

**Table 1:** Neonatal Characteristics.

| **Characteristic** | **Overall (N=320)** |
| --- | --- |
| **Place of birth** |  |
| Before arrival | 28 (8.8%) |
| Hospital | 292 (91.2%) |
| **Sex** |  |
| Female | 148 (46.2%) |
| Male | 172 (53.8%) |
| **Gestational Age** |  |
| Mean (SD) | 30.6 (2.95) |
| Range | 24 -35 |
| **Birthweight** |  |
| Mean (SD) | 1414.172 (411.361) |
| Range | 575.000 - 2435.000 |
| **Type of pregnancy** |  |
| Singleton | 219 (68.4%) |
| Twin | 85 (26.6%) |
| Triplets | 12 (3.8%) |
| Other (Quadruplet) | 4 (1.2%) |
| **Mode of delivery** |  |
| CS | 125 (39.1%) |
| SBD | 19 (5.9%) |
| SVD | 176 (55.0%) |
| **Admission Temperature** |  |
| Mean (SD) | 35.788 (0.982) |
| Range | 32.8 - 38.2 |

The mean age (SD) was 27(6.351) years and with a range of 15-43 years. The unemployed mothers were 179 (56.2%), 238 (74.4%) were married and 157(49.1%) attained up to secondary level. Only 3(0.9%) had a history of current cigarette smoking. (Table 2).

**Table 2:** Maternal demographic characteristics.

| **Variable** | **N** | **Freq (%)** |
| --- | --- | --- |
| **Age, n%** | **320** |  |
| Mean (SD) |  | 27.062 (6.351) |
| Range |  | 15 – 43 |
| **Occupation, n%** | **320** |  |
| Employed |  | 22 (6.9%) |
| Self-employed |  | 80 (25%) |
| Student |  | 38 (11.9%) |
| Unemployed |  | 180 (56.2%) |
| **Marital status, n%** | **320** |  |
| Divorced |  | 4 (1.2%) |
| Married |  | 238 (74.4%) |
| Single |  | 77 (24.1%) |
| Widowed |  | 1 (0.3%) |
| **Highest education level, n%** | **320** |  |
| None |  | 1 (0.3%) |
| Primary |  | 83 (25.9%) |
| Secondary |  | 157 (49.1%) |
| Tertiary |  | 79 (24.7%) |
| **Current smoking status** | **320** |  |
| No |  | 317 (99.1%) |
| Yes |  | 3 (0.9%) |

Among them, 162(50.8%) had a normal BMI, primiparas were 131(40.9%). A complete dose of antenatal steroids was administered to 78 (24.4%), Sixty-nine (21.6%) had a history of PPROM and among them 38(11.9%) were put on antibiotics and only 15(4.7%) presented with fevers. Majority, 314 (98.1%) were rhesus positive and those with blood group O+ were 131(40.9%). Among the maternal comorbidities, 3.4% tested positive for HIV and 100% of those that tested positive were started on ART. None had a viral load test done during pregnancy. One tested positive for syphilis and was put on treatment. 53(16.6%) had pre-eclampsia and only three (0.9%) had gestational diabetes mellitus. (Table 3)

**Table 3:** Maternal clinical characteristics.

|  |  |  |
| --- | --- | --- |
| **Parity** | 320 |  |
| Primipara |  | 131(40.9%) |
| Multipara |  | 189(59.1%) |
| **Antenatal steroids use** | 320 |  |
| No |  | 242 (75.6%) |
| Yes |  | 78 (24.4%) |
| **Rhesus** | 320 |  |
| -ve |  | 2(0.6%) |
| +ve |  | 314(98.1%) |
| Missing* |  | 4 (1.3%) |
| **BMI** | 320 |  |
| Underweight |  | 6 (1.9%) |
| Normal |  | 162 (50.6%) |
| Overweight |  | 120 (37.5%) |
| Obese |  | 32 (10.0%) |
| **HIV status** | 320 |  |
| Negative |  | 195 (60.9%) |
| Not tested |  | 114 (35.6%) |
| Positive |  | 11 (3.4%) |
| **Viral Load** | 11 |  |
| No |  | 11 (100.0%) |
| **On ARV** | 11 |  |
| Yes |  | 11 (100.0%) |
| **VDRL** | 320 |  |
| Negative |  | 298 (93.1%) |
| Not tested |  | 21 (6.6%) |
| Positive |  | 1 (0.3%) |
| **Treated** | 1 |  |
| Yes |  | 1 (100%) |
| **PPROM** | 320 |  |
| No |  | 251 (78.4%) |
| Yes |  | 69 (21.6%) |
| **Maternal Fever** | 320 |  |
| No |  | 305 (95.3%) |
| Yes |  | 15 (4.7%) |
| **Antibiotic use** | 320 |  |
| No |  | 282 (88.1%) |
| Yes |  | 38 (11.9%) |
| **Pre-eclampsia/Eclampsia** | 320 |  |
| No |  | 267 (83.4%) |
| Yes |  | 53 (16.6%) |
| **Chronic Hypertension** | 320 |  |
| No |  | 318 (99.4%) |
| Yes |  | 2 (0.6%) |
| **Pre-gestation DM** | 320 |  |
| No |  | 319 (99.7%) |
| Yes |  | 1 (0.3%) |
| **Gestational DM** | 320 |  |
| No |  | 317 (99.1%) |
| Yes |  | 3 (0.9%) |

Most of the neonates were put on non-invasive modalities. 265(82.8%) were started on low-flow nasal cannulae at 2 hours of life, 21(6.6%) were started on NCPAP, and 21(6.6%) on NIPPV. Seven (2.2%) were intubated and put on mechanical ventilation. At 72 hours, 110(34.5%) did not require any respiratory support, whereas 77(24.1%) were on low flow nasal canulae, 14(4.4%) on NCPAP, NIPPV were 65(20.4%) and 28(8.8%) were put on mechanical ventilation. Among the remaining 26 (7.8%) neonates, two were discharged and 24 died within 72 hours. (Table 4)

**Table 4:** Initial respiratory support.

| **Variable** | **N** | **Freq (%)** |
| --- | --- | --- |
| **Initial respirator support (2hours of life)** | **320** |  |
| Low flow nasal canulae |  | 265 (82.8%) |
| High flow Nasal canulae |  | 4 (1.2%) |
| NRM |  | 2 (0.6%) |
| NCPAP |  | 21 (6.6%) |
| NIPPV |  | 21 (6.6%) |
| Mechanical Vent |  | 7 (2.2%) |
| **Respiratory support at 72hours** | **320** |  |
| Low flow nasal canulae |  | 77 (24.1%) |
| NCPAP |  | 14 (4.4%) |
| NIPPV |  | 65 (20.4%) |
| Mechanical Vent |  | 28 (8.8%) |
| <sup>a</sup> Not on support |  | 136 (42.5%) |

<sup>a</sup> 110 – Room air, 24 – Died, 2 – discharged

Of the 313 put on noninvasive ventilation, 70(22.4%) [95%CI:17.95,27.47] had IRS failure characterized by stepping up respiratory support in the first 72 hours of life. 243(77.6%) had IRS success characterized by not stepping up respiratory support in the first 72 hours of life. The 7 neonates who had invasive ventilatory support as the IRS were excluded in this analysis. (Fig 3) We categorized the noninvasive IRS modalities into two groups as follows: Room air, Low/high flow nasal canulae and NRM as group one and NCPAP and NIPPV as a group two. This was statistically significant (p=<0.001) as group 1, 222(81.9%) had a higher IRS success rate compared to group 2, 21(50%). As shown on Table 5.

**Figure 3.**
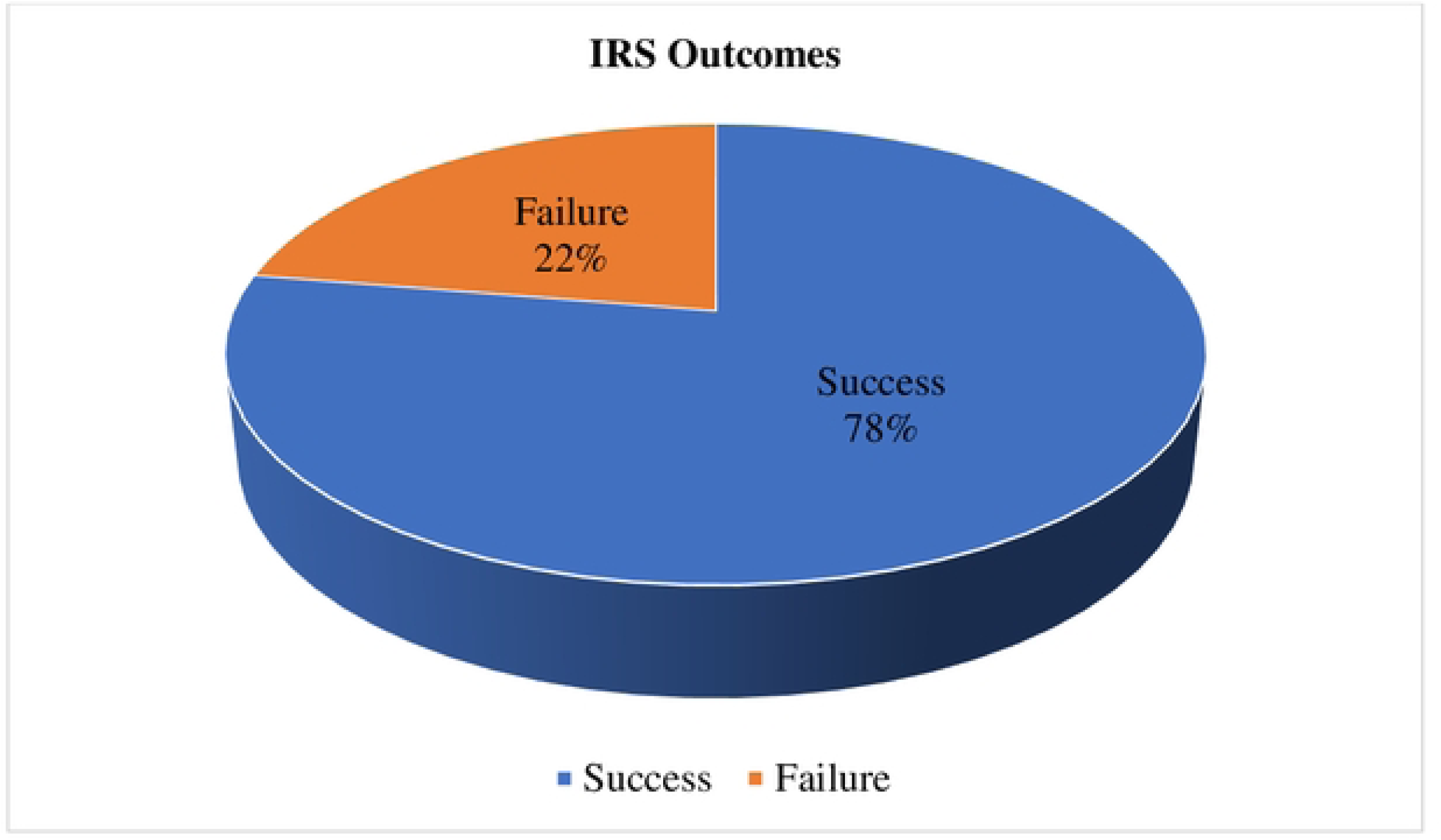

**Figure 4.**
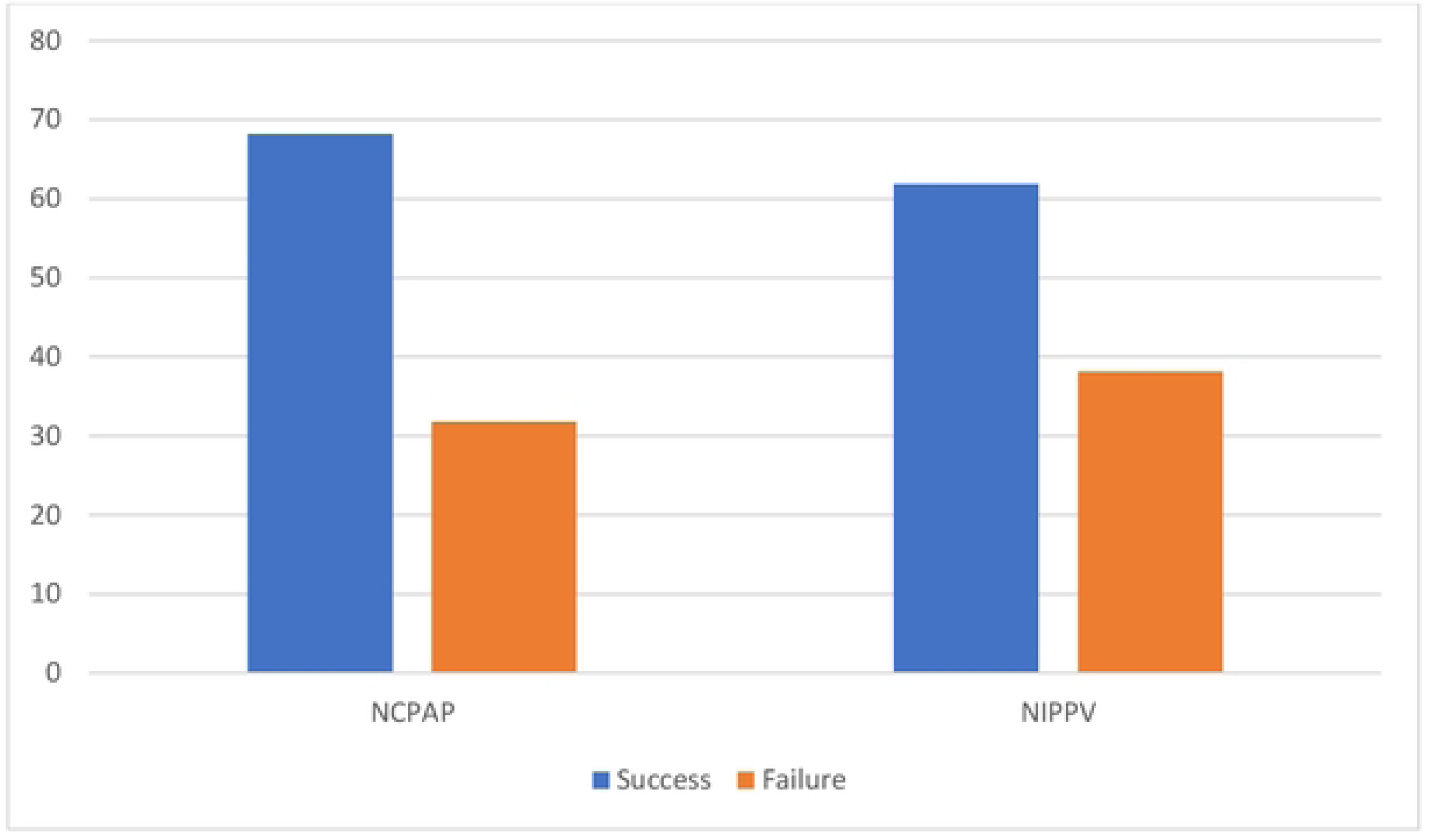

**Table 5:** IRS outcomes.

|  | Failure (N=70) | Success (N=243) | p value |
| --- | --- | --- | --- |
| **Technique** |  |  | **< 0.001** |
| Group 1 | 49 (18.1%) | 222 (81.9%) |  |
| Group 2 | 21 (50.0%) | 21 (50.0%) |  |

On further analysis of group 2, 13(61.9%) of neonates on NIPPV and 15(66.7%) on NCPAP did not need MV in the first 72 hours of life. (Fig 4)

On the bivariate analysis, the association between birth weight, respiratory distress based on modified Downes score, admission temperature, resuscitation at 15 minutes and IRS success was statistically significant. (p=<0.001).

Among the mothers, on bivariate analysis, level of education, parity and BMI were statistically significant (p=0.057, 0.001, 0.023). However, maternal age, occupation, smoking history, prenatal steroid use, Rhesus, Pre-eclampsia, and PPROM had no association with the IRS outcomes (p=0.529, 0.899, 0.349, 0.752, 0.446, 0.186, 0.794) respectively.

On multivariate analysis, the odds of IRS success were higher in neonates with a birthweight of > 1300g (AOR= 5.04;95%CI 1.81, 14.6), normal admission temperature (AOR=0.32;95%CI 0.12, 0.72), low modified Downes score (AOR=26.395%CI 3.37, 230), and neonates born to primiparas (AOR=2.81;95%CI: 1.42, 7.99) (p <0.05) Table 6 below.

**Table 6:** Multivariate Logistic regression of factors associated with IRS success.

| Characteristic | UO<br>R<sup>1</sup> | 95%<br>CI<sup>1</sup> | p-value | AOR<sup>1</sup> | 95% CI<sup>1</sup> | p-<br>valu<br>e |
| --- | --- | --- | --- | --- | --- | --- |
| **Birth weight** |  |  |  |  |  |  |
| <1000 g | 1 |  |  | 1 |  |  |
| 1000-1300g | 1.97 | 0.93,<br>4.19 | 0.076 | 1.23 | 0.45, 3.42 | 0.7 |
| >1300g | 10.1 | 4.62,<br>22.6 | <0.00<br>1 | 5.04 | 1.81, 14.6 | **0.002** |
| **Type of pregnancy** |  |  |  |  |  |  |
| Multiple | 1 |  |  | 1 |  |  |
| Singleton | 1.62 | 0.87,<br>3.19 | 0.15 | 2.25 | 0.95, 5.62 | 0.072 |
| **Mode of Delivery** |  |  |  |  |  |  |
| CS | 1 |  |  | 1 |  |  |
| SBD/SVD | 1.55 | 0.90,<br>2.70 | 0.12 | 1.32 | 0.57, 3.08 | 0.5 |
| **Temperature** |  |  |  |  |  |  |
| Normal | 1 |  |  | 1 |  |  |
| Mild | 1.30 | 0.51,<br>3.4 | 0.6 | 1.26 | 0.41,3.82 | 0.7 |
| Moderate | 0.26 | 0.12,<br>0.51 | <**0.00**<br>**1** | 0.32 | 0.12, 0.82 | **0.003** |
| **Modified Downes**<br>**score** |  |  |  |  |  |  |
| Impending arrest | 1 |  |  | 1 |  |  |
| Moderate | 1.6 | 0.29,<br>8.86 | 0.6 | 1.78 | 0.27, 12.5 | 0.5 |
| Mild | 28.2 | 4.34,<br>193 | <0.00<br>1 | 26.3 | 3.37, 230 | **0.002** |
| **Resuscitation**<br>**15minutes** |  |  |  |  |  |  |
| None | 1 |  |  | 1 |  |  |
| Oxygen | 0.63 | 0.31,<br>1.20 | 0.2 | 0.86 | 0.33, 2.15 | 0.7 |
| **Level of Education** |  |  |  |  |  |  |
| Above Primary | 1 |  |  | 1 |  |  |
| Primary and below | 0.58 | 0.33,<br>1.03 | 0.059 | 0.56 | 0.25, 1.22 | 0.14 |
| **BMI** |  |  |  |  |  |  |
| Normal/underweig<br>ht | 1 |  |  | 1 |  |  |
| Obese/overweight | 0.83 | 0.48,<br>1.41 | 0.5 | 1.3 | 0.62, 2.78 | 0.5 |
| **Parity** |  |  |  |  |  |  |
| Multipara | 1 |  |  | 1 |  |  |
| Primipara | 2.79 | 1.54,<br>5.29 | 0.001 | 3.28 | 1.42, 7.99 | **0.007** |
| **Pre-eclampsia** |  |  |  |  |  |  |
| No | 1 |  |  | 1 |  |  |
| Yes | 1.71 | 0.80,<br>4.10 | 0.2 | 1.36 | 0.45, 4.42 | 0.6 |

## Discussion

### Initial respiratory support outcomes

We sought to determine the initial respiratory support outcomes of RDS among the preterm neonates with RDS. Most neonates on NIV had IRS success (77.6% vs. 22.4%). This mirrors findings in the study by Fallahi et al which had a success rate of 62.2% in the (Non-Invasive group): Oxyhood [11 (13.40%) neonates], Room Air [14 (17.10%) neonates], NCPAP [19 (23.20%) neonates], and non-invasive positive pressure ventilation [NIPPV, 38 (46.30%) neonates (18)]. This was also similar to VENTIS study that reported a global incidence of NIV failure of 15.6% (success; 84.4%) (19). However, these findings were dissimilar to the study by Afjeh et al, where the noninvasive respiratory initial respiratory support (RA, oxygen via prongs, NCPAP/NIPPV) was successful in 147(41.7%) but failed in 175 (54.3%) neonates. (15). This could be attributed to a difference in study participants of VLBW neonates whereas our study had neonates ranging 575g to 2435g.

Group 1(RA, Low/high nasal canulae/NRM) had a success of 222 (81.9%). This is similar to those of Afjeh et al group 1(RA, oxygen therapy) who reported IRS success of 77.7%. (15) Fallahi et al had similar success rates in the noninvasive group which included room air and oxyhood were 12 (85.70%) and 11 (100%). (18)

In this current study, patients whose IRS was NCPAP, (success; 66.7% vs. failure; 33.3%). Prior studies focused solely on the CPAP technique and found a higher percentage of success. Dargaville et al. reported a success rate of 78% with 65(22%) failing CPAP as the IRS (12) Afjeh et al. reported NCPAP success; 80.4% vs. failure; 19.6%.(15). Ammari and colleagues established NCPAP success;76% vs. failure; 24% (20). Contrary to this study, some studies had lower IRS success rates. Fuchs et al reported NCPAP success rate of 68/140 (49%). (21) Correspondingly, another study by Dunn and colleagues, reported success in 48% (22). This difference can be attributed to the fact that both study populations were composed of neonates below 29 weeks.

In this present study, neonates whose IRS was NIPPV, only 8(38.1%) out of 21 neonates started on NIPPV failed (NIPPV success; 61.9%% vs. failure; 38.1%). Likewise, Afjeh et al study, NIPPV success; 60.1%% vs. failure; 39.9%. (15). In other comparable findings, Manzar et al reported NIPPV reduced the need for intubation and mechanical ventilation in 81% of the cases (Manzar et al., 2004), Kugelman et al reported NIPPV success of 75%(23) and Meneses et al, 75% neonates on NIPPV were weaned off to NCPAP in the first 72 hours and did not need MV. (24) Contrary to this, Fallahi et al. reported NIPPV success rate of 16(42.10%) (18) However since NIPPV was mostly used as a rescue therapy for the neonates on NCPAP in the first 72 hours and some neonates died when still undergoing treatment on NIPPV, as a result, individual analysis of NIPPV failure was not practicable.

### Variables associated with the non-invasive ventilation success

Neonates weighing more than 1300g had a higher chance of success than those below 1000g(p=<0.001). This finding is similar to Afjeh et al who found odds of success for neonates above 1000g was 3.3 times (AOR =3.34);95%CI 1.64, 6.82) that of neonates below 1000g. (p=<0.001). (15) In a more recent study by Yaser et al birth weight ≤ 1200g was the only factor found to be associated with CPAP failure (OR 2.05 CI 1.07– 3.92, p 0.029). (25). In contrast, Fallahi et al showed no association between birthweight as a predictor for success or failure in selecting IRS (AOR=1.14;95%CI 0.48, 2.71, p=0.752). (18). This difference can be attributed to the fact that Fallahi and colleagues’ study compared the success of IRS between the invasive and noninvasive groups whereas our study only assessed success in the noninvasive group.

In this study, neonates with mild respiratory distress based on modified Downes score had a higher odds of IRS success than those with impending respiratory distress (p=<0.002). This finding is similar to Permatahati et al study where 57.5% of infants with initial Downes score >3 experienced nasal CPAP failure (*p* =0.035) by multivariate analysis. Downes score >3 could predict CPAP failure 2.68 times more than Downes score <3 (AOR=2.68;95%CI 1.07,6.72).(26) Likewise, Koti et al study also established a Downes score of >7 was associated with CPAP failure (p=0.003).(27). In this study, the odds of IRS success among neonates with moderate hypothermia was less than that of neonates with normal admission temperatures (p=<0.003). This study finding is attributed to the normalcy of physiology of hypothermia that leads to increased oxygen consumption and pulmonary vasoconstriction, increased respiratory distress and reduced surfactant release. (28) Similar to our study, Carns et al study on the impact of hypothermia on outcomes for preterm infants with RDS treated with CPAP and nasal oxygen in Malawi concluded that hypothermia of below 35.8°C had poor outcomes in neonates with RDS managed with CPAP or nasal oxygen as none of them survived. In contrast, neonates treated with CPAP with more than 35.8°C the survival rate was 100% and 36.4% for the 11 neonates treated with nasal oxygen (p<0.001). (29)

Parity was associated with IRS outcomes. The odds of neonates born by primiparas of having a successful IRS was 3.2 times that of neonates who were born by multiparas (p=0.001). Contrast to this finding was the study by Bacha et al where parity was not significant in determining the outcome of RDS. (30) This difference is because Bacha and colleagues did not associate parity and IRS outcomes which was the case in ours. No other studies have compared parity to IRS outcomes.

### Conclusions and recommendation

In this study, noninvasive initial respiratory support (IRS) success occurred in majority of the neonates 243(77.6%). Group 1 had a higher IRS success rate compared to Group 2. Primiparity, birthweight of more than 1300g, normal to mild hypothermia at admission, and low modified Downes score were associated with IRS success. With the foregoing, there is a need to adopt a modified Downes score within the existing protocols as a monitoring tool for neonates on NIV and prevention of hypothermia to improve IRS success. Further multicenter RCT studies are recommended to further analyze predictive variables for early identification of neonates likely to fail NIV IRS to allow specific therapeutic interventions aimed at them.

## Data Availability

All relevant data are within the manuscript and its Supporting Information files.

